# Pre-diagnosis blood DNA methylation profiling of twin pairs discordant for breast cancer points to the importance of environmental risk

**DOI:** 10.1101/2023.08.15.23293985

**Authors:** Hannes Frederik Bode, Liang He, Jacob V.B. Hjelmborg, Jaakko Kaprio, Miina Ollikainen

**Affiliations:** Institute for Molecular Medicine Finland, HiLIFE, University of Helsinki, Tukholmankatu 8, 00290 Helsinki, Finland; Department of Public Health, University of Southern Denmark, Campuses 55, 5230 Odense M, Denmark; Minerva Foundation Institute for Medical Research, Tukholmankatu 8, 00290 Helsinki, Finland

**Keywords:** DNA methylation, breast cancer, survival analysis, monozygotic twins, dizygotic twins

## Abstract

**Background:** Assessment of breast cancer (BC) risk generally relies on mammography, family history, reproductive history, and genotyping of major mutations. However, assessing the impact of environmental factors, such as lifestyle, health related behavior or external exposures, is still challenging. DNA methylation (DNAm), capturing both genetic and environmental effects, presents a promising opportunity. Previous studies have identified associations and predicted the risk of BC using DNAm in blood, however, these studies did not distinguish between genetic and environmental contributions to these DNAm sites. In this study, associations between DNAm and BC are assessed using paired twin models, which control for shared genetic and environmental effects, allowing testing for associations between DNAm and non-shared environmental exposures and behavior.

**Results:** Pre-diagnosis blood samples of 32 monozygotic (MZ) and 76 dizygotic (DZ) female twin pairs discordant for BC were collected at the mean age of 56.0 years, with the mean age at diagnosis 66.8 and censoring 75.2 years. We identified 212 CpGs (p<6.4*10^−8^) and 15 DMRs associated with BC risk across all pairs using paired Cox proportional hazard models. All but one of the BC risk associated CpGs were hypomethylated and 198/212 CpGs had their DNAm associated with BC risk independent of genetic effects. According to previous literature, at least five of the top CpGs were related to estrogen signaling. Following a comprehensive two-sample Mendelian Randomization analysis, we found evidence supporting a dual causal impact of DNAm at cg20145695 (gene body of *NXN*, rs480351) with increased risk for estrogen-receptor positive BC and decreased risk for estrogen-receptor negative BC.

**Conclusion:** While causal effects of DNAm on BC risk are rare, most of the identified CpGs associated with the risk of BC appear to be independent of genetic effects. This suggests that DNAm could serve as a valuable biomarker for environmental risk factors for BC, and may offer potential benefits as a complementary tool to current risk assessment procedures.

## Background

Breast cancer (BC) risk assessment tools rely on physiological or genetic screening methods such as mammography, family history and information on reproductive history. In addition to assessment of major mutations, polygenic risk scores are increasingly used to assess overall genetic risk. However, accurately quantifying the impact of environmental factors, including health-related behaviors and occupational exposures, can be challenging, and these factors are often not included in the risk assessment models. In recent years, DNA methylation (DNAm) has emerged as a promising biomarker for BC risk assessment.

BC has been linked to DNAm in blood, as evidenced by specific DNAm sites [1–19] and overall average DNAm levels [20] associating with BC. In addition, the studies conducted by Kresovich [2021], Xiong [2022] and Chung [2023] and colleagues [4,16,21] have shown that blood-derived DNAm can be used to predict an individual’s risk of developing BC. This presents an opportunity for blood-derived DNAm to serve as a complementary measure to current standard BC risk assessment tools [22,23]. However, these predictors, as well as earlier work on DNAm and overall BC risk [2,4,6,14,16,17,19] have not differentiated between genetic and environmental risk factors for BC.

Health-related behaviors and environmental exposures are major factors that can increase the risk of BC [24], and many of these factors have been shown to affect DNAm as well; e.g. alcohol use [25–27], obesity [28,29], physical inactivity [30], hormonal exposure [31], and reproduction-related factors [29,31–33]. In addition, genetic variants, including those linked to BC risk, may affect DNAm.

Twin pairs discordant for disease provide a valuable approach for investigating the impact of environmental factors. Within monozygotic (MZ) twin pairs the genetic components are fully controlled for as the co-twins are genetically identical at the germline. Genetic effects are also partially controlled for in the within-pair comparison of dizygotic (DZ) twin pairs, in which the twins share on average 50% of their segregating genetic background. Moreover, the twin design effectively controls for age and all shared environmental influences, especially in early life, regardless of whether they are DZ or MZ twin pairs. Although the discordant MZ twin pair design does not capture *de-novo* mutations it dramatically boosts the statistical power and allows robust investigation of the relationship between environmental BC risk and DNAm, while completely controlling for genetic confounding.

In this study, the objective was to investigate the potential of DNAm as a biomarker for environmental BC risk, and further to assess causality. To accomplish this, we leveraged a cohort of BC discordant twin pairs from the Finnish Twin Cohort with DNAm data collected prior to BC diagnosis. Then, we aimed to validate the findings by examining an independent BC discordant twin pair dataset from the Danish Twin Study. Finally, we used two-sample Mendelian Randomization analysis to find further evidence for causality.

## Methods

### The Finnish Twin Cohort

The Finnish Twin Cohort (FTC), consisting of twins from like-sexed pairs born before 1958, was established in 1975, recruitment was completed by May 1, 1976, and the follow-up period lasted until December 31, 2018. To obtain cancer diagnosis data during the study period, the FTC was linked to the Finnish Cancer registry. Information on death and emigration was obtained from the Finnish population registry. Starting in the 1990s, blood samples were collected from a subset of individuals and DNA was extracted and stored in the Biobank of the Finnish Institute for Health and Welfare. DNAm data was subsequently generated from these samples.

A group of 108 pairs of female twins who showed discordance for BC at the end of the follow-up period were selected from among all pairs with DNA from the FTC. Of them, 32 pairs were MZ, and 76 pairs were DZ (Table 1). Among the cases, BC was either their first or only cancer diagnosis, while the controls remained cancer-free during the follow-up. The follow-up period was considered to end for cases at the time of diagnosis and for controls either at death (n=18) or latest at the end of the study in 2018 (n=90).

**Table 1:**
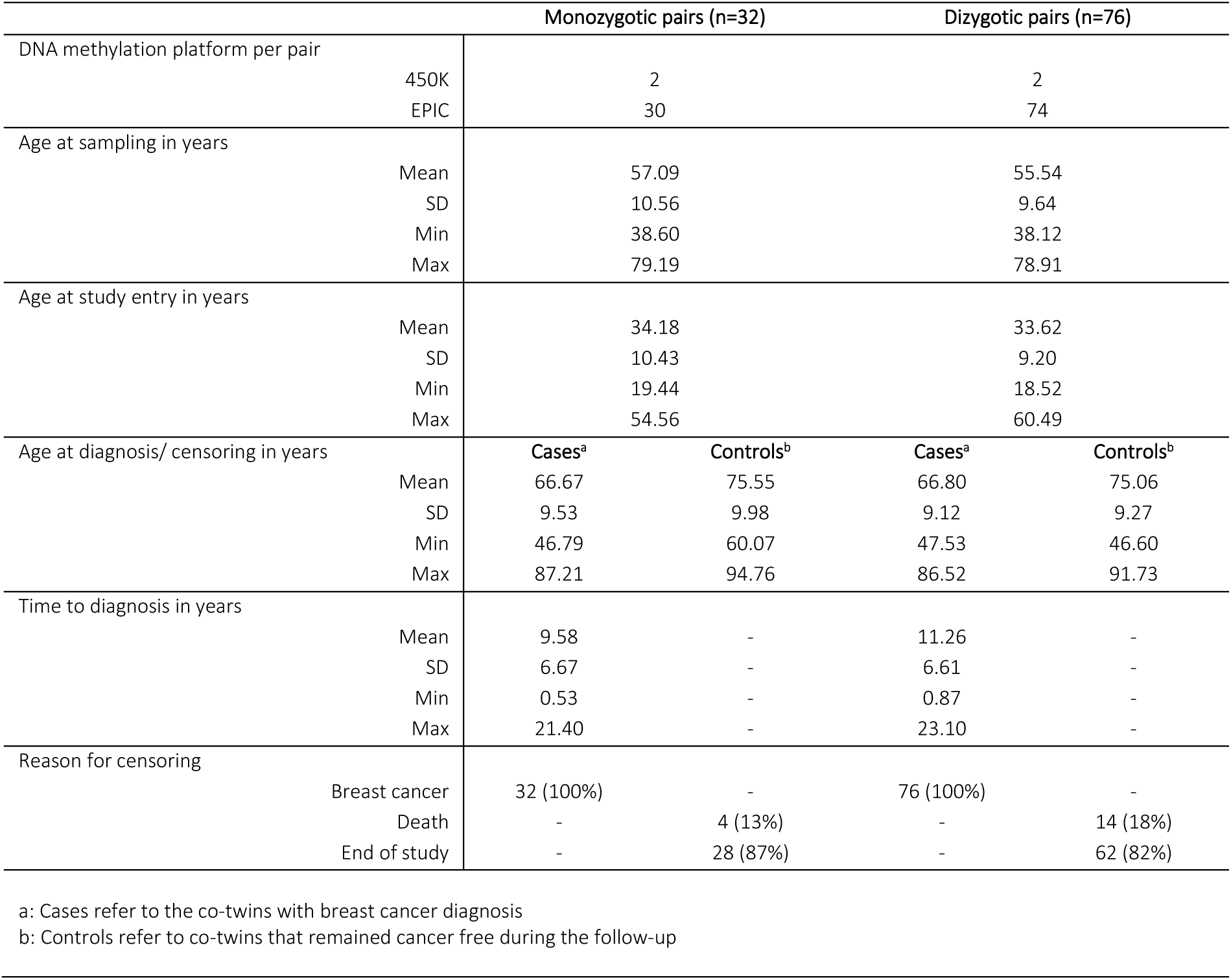
Description of Finnish Twin Cohort variables used in the survival models.

Data on epidemiological risk factors for BC were obtained from a health-related questionnaire collected in 1975 (Table 2), while information on the number of children and age at first birth was obtained from the Finnish Population Register for individuals born 1950 onwards (Table 2) [34]. The association between these variables and BC discordance was examined using conditional logistic regression for MZ and DZ twin pairs together (Table 2).

**Table 2:**
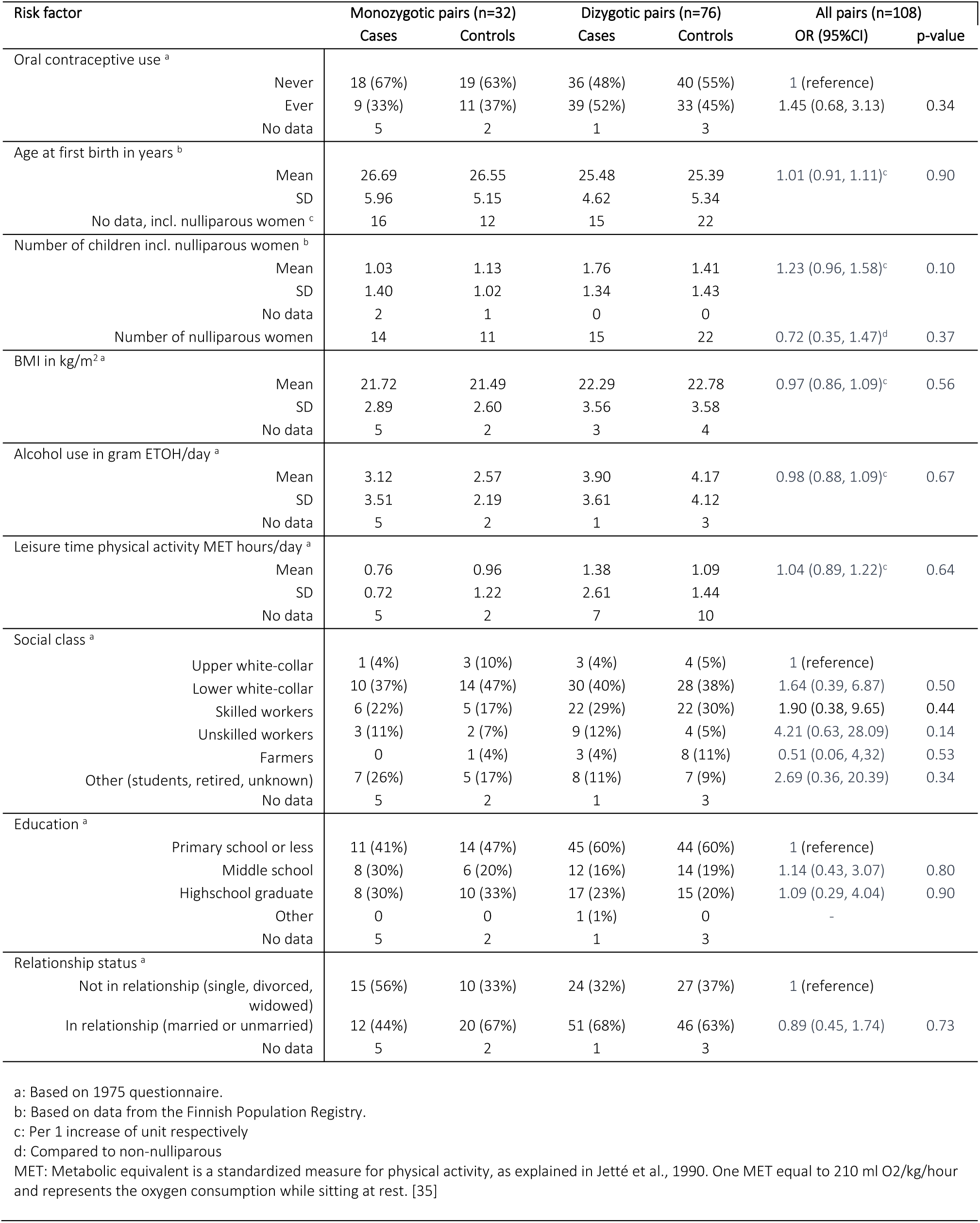
Description and Comparison of the available breast cancer risk factors for FTC data.

### The Danish Twin Study

The Danish sample is selected from two Danish twin cohorts in the Danish Twin Study (DTS), including the Longitudinal Study of Aging in Danish Twins (LSADT) study and the Middle Age Danish Twins (MADT) study. Details about these twin cohorts are described previously in [36,37]. Only included those twins that have both DNAm (1180 samples) and the information about BC diagnosis were included, which was retrieved through the link between the twin registry and the cancer registry in the NorTwinCan database [38]. Eighty-six twins in LSADT were measured twice in 1997 and 2007, respectively, for their DNAm and only the early measurement in 1997 were included to increase the sample size for the analysis of pre-diagnosis DNAm. Among these twins, 11 twin pairs (eight MZ and three DZ pairs) that are discordant for BC and of whom the methylation was measured prior to the diagnosis were included in the analysis. The end of follow-up was defined the same way as for the Finnish dataset.

### DNA methylation data

Among the 108 pairs from the FTC, DNAm was measured for four pairs using the Illumina Infinium HumanMethylation450 (450K) platform and for 104 pairs the Illumina Infinium MethylationEPIC (EPIC) platform (Illumina, San Diego, CA, USA), (Table 1). Twins in a pair shared the same platform technology and were sampled and processed at the same time. Preprocessing of the DNAm data was done using the *meffil* R package [39]. Sample quality was assessed based on the following criteria: (1) mean difference between X and Y (technical noise in female samples) chromosome signals was less than −2, (2) mean methylated signal did not deviate from the regression line (mean methylated signal linear regressed over mean unmethylated signal) by more than three standard deviations, (3) sample was not an outlier based on Illumina control probes, (4) percentage of probes with only background signal was less than 20, and (5) percentage of probes with less than three beads was less than 20. All samples that met all the above criteria passed the quality control. In addition, ambiguous mapping and poor-quality probes based on Zhou et al. (2017, [40]) and probes binding to sex chromosomes were removed to address ambiguity of the DNAm signal based on X-chromosome inactivation [41,42]. Following the quality control, the DNAm data underwent preprocessing in two separate batches due to use of two different types of microarray platforms, EPIC and 450K.

The preprocessing was performed by functional normalization using the first 15 principal components of the control probes, to eliminate unwanted technical variation using the *meffil* R package [39]. Additionally, to reduce probe bias, beta-mixture quantile normalization [43] using the *wateRmelon* R package [44] was applied. Next, the CpG probes exclusively present on the EPIC platform were merged with the set of probes common between the EPIC and 450k platforms. Finally, the beta values were scaled based on the standard deviation of each CpG probe across all samples. For the annotation of the CpG sites the latest version of the *Illumina Infinium Methylation EPIC manifest v1.0 B5* was used (Illumina, San Diego, CA, USA).

After performing quality control and preprocessing, a total of 52333 probes were removed due to their insufficient quality, and 9918 probes were removed due to their binding to sex chromosomes. A final set of 778861 probes were retained, consisting of 336849 probes (43%) shared by the 450K and EPIC data and 442012 probes exclusive to the EPIC platform.

In the DTS data, DNAm was measured from the buffy coat samples stored at −80°C in 24 hours after the blood was collected using the 450K platform (Illumina, San Diego, CA, USA). Quality control for sample and probe exclusion were conducted with the *MethylAid* [45] and *Minfi* [46] R packages. More detailed steps of quality control and criteria for excluding samples and probes are described in the previous study [47]. After further excluding any CpG sites that had a missing rate of >10% across the whole 1180 samples, 451471 CpG sites remained in the survival analysis.

### Survival modelling

To investigate the association between DNAm and BC risk, a survival analysis was conducted using Cox proportional hazard regression models in the R package *survival* [48]. To ensure that all CpG sites meet the model assumptions, the proportional hazard assumption for the within-pair difference in methylation beta value was examined by Schoenfeld residuals and tested using the *cox.zph()* function for the significant CpG sites. For the analysis, four different Cox Proportional Hazard models were applied. The equations for all four models are the same, only the cohorts tested with these models different, e.g. MZ twin pairs, DZ twin pairs, MZ and DZ twin pairs together, and twin pairs from the DTS (Equation 1).

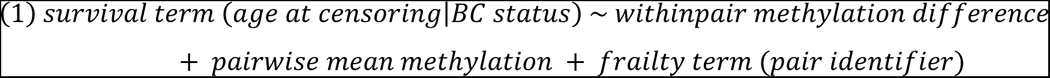

The survival term in this model refers to the survival outcome observed during the follow-up period. To adjust for the methylation levels specific to each twin pair, the mean beta values for each pair were included as an explanatory variable. The pair identifier is considered as a random effect. Additionally, based on the study design the pairs were matched on potential confounding variables, such as chip platform, age at entry, age at sampling, sex, and early life environment.

The discovery cohort involved 108 twin pairs that were sampled before the onset of BC diagnosis (Model 1). Using Bonferroni method for multiple testing correction, p-values < 6.4*10^−8^ were considered significant. The statistically significant CpG sites were followed up in zygosity-specific analyses of MZ pairs (n=32, Model 2) and DZ pairs (n=76, Model 3), to assess the role of genetic vs environmental effects on the significant methylation sites. CpG sites with the same effect direction compared with Model 1 and p < 0.05 were considered as validated.

To replicate the findings in an independent twin sample, the same survival model was fitted using the DTS data of 11 twin pairs discordant for BC (Model 2R). The CpG sites were considered as replicated when the direction of the effect was the same as in Model 1 and p-value < 0.05.

### Sensitivity analyses

To evaluate the possible effect of unmeasured confounding variables on the association between DNAm and survival outcomes, E-values were computed for each significant CpG site using the HR of such in the EValue R package [49,50]. E-values for HR > 1 follow equation 2 [49]:

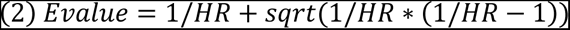

E-values for HR < 1 follow equation 3 [49]:

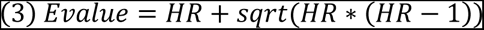

### DMR analysis

Differentially methylated regions (DMR) were identified using the *ipDMR()* function [51] of the *ENmix* R-package [52]. Hereby neighboring CpG sites within a maximum distance of 500 base pairs were identified, and their combined p-values were calculated using the p-values derived from Model 1. Using a Benjamini-Hochberg FDR < 0.001, all significant pairs of CpG sites were selected. These significant CpG pairs were then merged into broader regions using an approach like single linkage clustering, where neighboring CpG sites within a maximum distance of 500 base pairs were linked together. The association between each broader region and the survival outcome was then calculated by summarizing the associations of all CpG sites present in such region, based on the individual CpG sites p-values derived from Model 1. A DMR was considered significant if it contained three or more CpG sites with unidirectional methylation association and had a Benjamini-Hochberg FDR < 0.001 combined across all CpG sites.

### Mendelian Randomization

To test whether DNAm at the identified CpG sites is causal for BC risk a two-sample Mendelian randomization (MR) analysis was performed. We established valid genetic instrumental variables (IVs) for the BC–associated DNAm from Model 1, with the following criteria: The identified SNP to be used as IV 1) associates with DNAm at give CpG site of interest (associated with BC under Model 1) with genome-wide significance, 2) does not directly associate with breast cancer, and 3) does not associate with confounders of BC risk or DNAm. First, for the 212 BC–associated CpG sites (Model 1) 18 cis-meQTL SNPs with genome wide significance (p < 6.9*10^−8^) were identified using the *MeQTL EPIC Database* (https://epicmeqtl.kcl.ac.uk/) [53].

Second, summary statistics from two BC genome-wide association studies (GWAS) were used to eliminate any IVs that directly associate with BC. The first set of summary statistics were derived from the IEU GWAS project (https://gwas.mrcieu.ac.uk/) [54], comprising 212402 female individuals (6.53% cases) from the UK Biobank (UKBB) made publicly available by the University of Bristol under a non–commercial government license [55], and the other set was from the FinnGen study release R9 [64] under access-rights received on 26th September 2023, encompassing 222080 female individuals (9.27% cases). Among these, GWAS summary statistics were available for 13 and 12 out of the 18 meQTL SNPs, respectively, and none of these exhibited statistically significant genome wide association with BC. Additionally, none of the 12 meQTL SNPs available in the FinnGen estrogen receptor positive (ER+) BC GWAS on 213307 female individuals (5.66% cases) and estrogen receptor negative (ER-) BC GWAS on 209695 female individuals (4.03% cases) had genome-wide significant association with these BC subtypes.

Third, to eliminate IVs that associate with potential confounders, a phenome-wide association analysis was performed using the *PhenoScanner V2* (www.phenoscanner.medschl.cam.ac.uk) [56]. No association with any of the available trait or disease was observed for the 13 SNPs, however, one SNPs associated with the level of monocytes and granulocytes, a potential confounder of DNAm, resulting to 12 suitable IVs.

To assess the relationship between exposure (DNAm at the given CpG site) and the outcome (risk for BC), Wald’s ratio testing was employed for each individual SNP (Supplementary Figure 1), which estimated the causal effect of DNAm on BC risk at the given CpG site. Subsequently, Egger MR analysis was performed to test pleotropic effects on BC risk across the identified IVs. To test for the directionality from DNAm to BC (BC overall, ER+ and ER-BC) risk the MR Steiger test was performed [57]. An association was considered significant if both the Wald’s ratio test and the MR Steiger had nominal p-values < 0.05 and the MR Steiger test indicated directionality from DNAm on BC risk. The MR analysis was carried out using the R package *TwoSampleMR* [58].

## Results

### Discovery

Altogether 108 BC discordant FTC twin pairs (n= 32 MZ and n=76 DZ pairs) sampled prior to BC diagnosis were used as the discovery sample (Table 1 and 2). The average age of study entry was 33.8 years (SD = 9.5 years) and the average age for blood sample collection was 56.0 years (SD = 9.9 years). For cases, the age at diagnosis was on average 66.8 years (SD = 6.6 years), and for controls, the end of follow-up was at an average age of 75.2 years (SD = 9.5 years). The mean time between blood sampling and diagnosis was 10.8 years (SD = 6.7). The association between these variables and BC discordance was examined using conditional logistic regression for MZ and DZ twin pairs together. None of these variables showed a significant association with BC discordance (Table 2).

The first survival model (Model 1) included MZ and DZ pairs, and matched for familial effects. This discovery analysis resulted in 212 DNAm sites significantly associated (p < 6.4*10^−8^) with future BC (Figure 1A, and Supplementary Table 1). Among these CpG sites, all except one (cg00550725, in *FAM82B*, more commonly known as *RMDN1*) showed negative association, as indicated by Hazard Ratio (HR) below one, implying that lower DNAm levels associated with higher hazards of BC. The BC–associated hypomethylated CpG sites had HRs ranging from 0.01 to 0.49, while the hypermethylated CpG site had an HR of 3.07. *TDRD1* was the only gene with two significant BC–associated CpG sites (cg14779973 and cg27547703).

**Figure 1:**
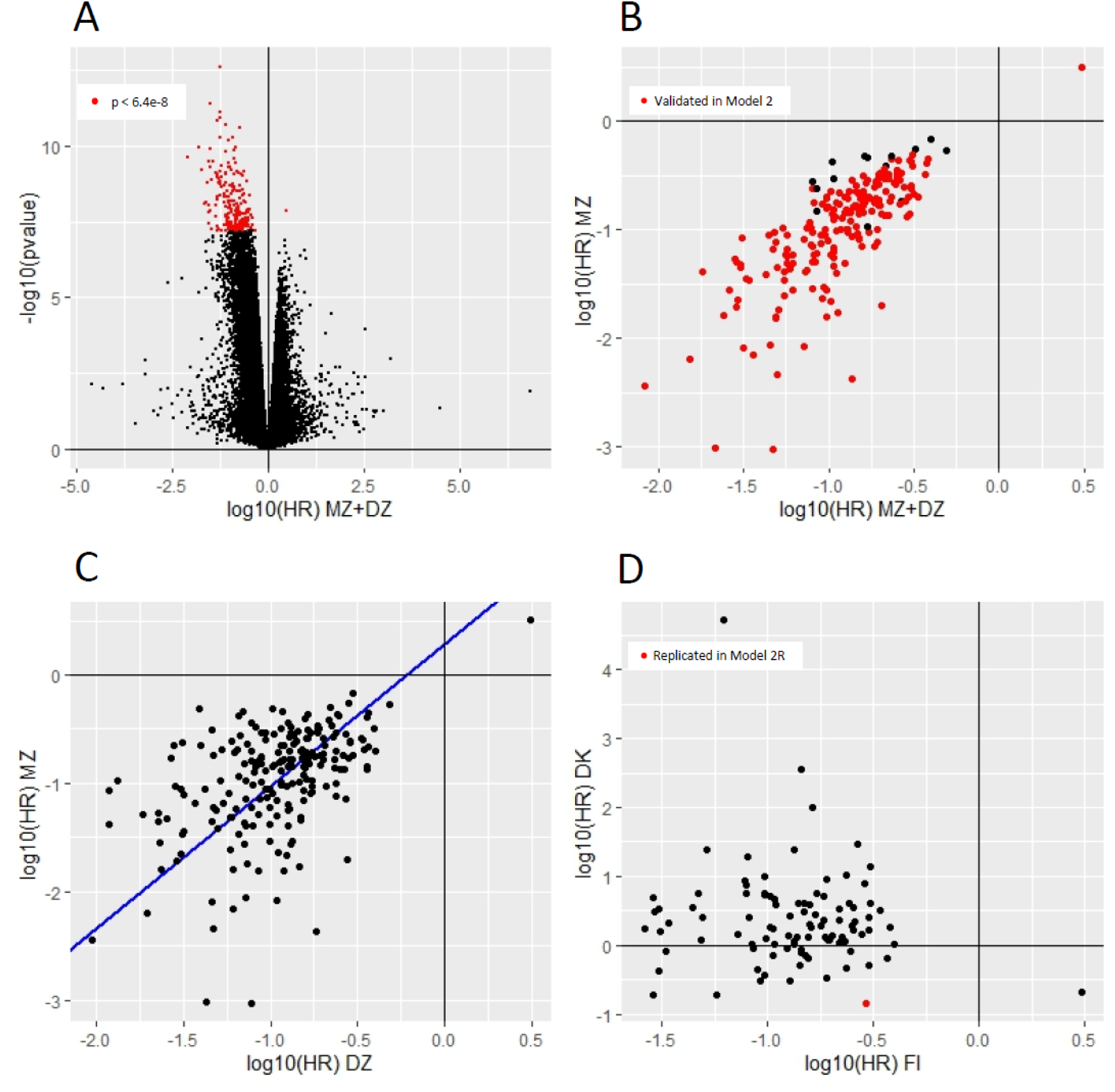
Results on the survival modeling for individual CpG sites associated with breast cancer; A) Volcano plot of Model 1 (MZ+DZ) with CpG sites significantly associated with breast cancer marked in red; B) Comparison between Model 1 (MZ+DZ) and Model 2 (MZ) with significant CpG sites from Model 1 validated in Model 2 marked in red; C) Comparison between Model 2 (MZ) and Model 3 (DZ) with a regression line in blue (regression coefficient = 1.31, p=0.001); D) Comparison between Model 1 (MZ+DZ, Finnish data) and Model 2R (MZ+DZ, Danish data) with replicated CpG site in red.

A sensitivity analysis was conducted by calculating E-values for the 212 significant CpG sites in Model 1 (Supplementary Table 1). The E-values were high for all CpG sites, indicating that unknown or unmeasured covariates are unlikely to account for the association between DNA methylation at these CpG sites and BC.

In addition to the 212 individual CpG sites, 15 DMRs were significantly associated with BC in Model 1 (Supplementary Table 2). Among these, three DMRs (in genes *SCMH1*, *PXDNL* and *GNAS*, all relevant for BC biology) contain CpG sites that were also significant as single hits in Model 1. Out of the 15 DMRs, 14 have lower DNAm (average HR<1) in the BC diagnosed twin compared with their co-twin.

### Validation

To explore whether the 212 BC–associated CpG sites are likely due to environmental effects, we performed within-pair analysis including only MZ twin pairs, which rules out the potential genetic confounding in the observed associations (Model 2). Altogether 198 CpG sites (93%) showed the same effect direction and met nominal significance (p < 0.05) for association with BC (Supplementary Table 1, Figure 1B).

We next assessed the associations in Model 3 containing DZ twin pairs only, which resulted in all the 212 CpG sites having the same effect direction as in Model 1 and 2, and meeting nominal significance (p < 0.05) for the association with BC (Supplementary Table 1). However, on average the 212 CpG sites had higher effect sizes in the Model 2 compared with Model 3 (beta = 1.31, p = 0.001), suggesting that a higher level of genetic matching results in higher effect size (Figure 1C).

### Replication with the Danish Twin study

We aimed at replicating the findings from the FTC in the DTS containing 11 twin pairs discordant for breast cancer (eight MZ and three DZ pairs). The mean age at the diagnosis of these 11 twin pairs was 78.1 (SD=9.6 years) and the mean age at the DNAm profiling was 71.3 (SD=6.6 years). The DTS datasets included 98 out of the 212 CpG sites identified in the discovery analysis performed in FTC Model 1. The remaining sites were not present in the DTS data, which was generated using the smaller 450K methylation platform. Out of these 98 CpG sites 22 had the same effect direction in the DTS (Model2R) as in the FTC data (Model 1 and Model 2). However, only one of these CpG sites (cg16376218, within *SLC25A39*) met nominal significance (p < 0.05) for the association with BC (Supplementary Table 1, Figure 1D).

### Mendelian randomization

Our study design, DNAm measured before BC diagnosis and the within-pair comparisons ruling out confounding by shared genetic and environmental effects, suggests that the observed BC–associated DNAm patterns precede BC. We next aimed at assessing if there is additional evidence for direct causation by performing a two-sample MR analysis. Altogether 12 genome-wide significant meQTLs met the criteria (see Methods) for IVs and were used in the two-sample MR to test for causality for BC in general, and for ER+ and ER-subtypes. The MR analysis was performed using Wald’s Ratio testing for individual IVs. Egger regression analysis across all available IVs revealed no pleiotropy, and the Steiger MR test showed that directionality went from DNAm to BC risk for all IVs.

No causal effects were observed for BC risk in general for the 12 CpG sites (Supplementary Table 3). Using the 11 out of 12 IVs available for ER+ and ER-BC (Supplementary Table 3), DNAm at cg20145695 (*NXN*, rs480351) only showed significant causal effect. Higher methylation increased the risk for ER+ BC (OR_ER+_= 1.51 (CI95 = 1.02-2.24), p = 0.04) and decreased the risk for ER-BC (OR_ER-_= 0.72 (CI95 = 0.52-0.98), p = 0.04), showing that DNAm at this site has effects opposing each other depending on the BC subtype. (Supplementary Table 3).

## Discussion

Here we studied the association between BC risk and DNAm in blood samples taken prior to cancer diagnosis. We found that 212 CpG sites and 15 DMRs in blood were associated with the risk of BC using a discordant twin study design, matched for familial confounders. Among the 15 DMRs we identified, three contain CpG sites that also associate individually with BC risk. These CpG sites are located in the genes *SCMH1*, *PXDNL*, and *GNAS*. We validated the majority of the BC risk associated CpG sites (198/212) within MZ pairs, and with significantly higher effect sizes than within DZ pairs, suggesting that these 198 CpG sites associate with BC independent of genetic factors and are likely attributed to environmental BC risk. Only one of these significant sites (cg16376218 in *SLC25A39*) replicated in the Danish Twin Study.

Our study validated one previously observed BC–associated CpG site (cg21769444, *NUDT3*) [18]. *NUDT3* associated long noncoding RNA, NUDT3-AS4, appears to promote cell growth in BC. It acts as a sponge for microRNA miR-99s, competing with AKT1/mTOR mRNAs for binding. This competition prevents miR-99s from degrading AKT1/mTOR mRNAs, leading to increased expression of AKT1 and mTOR proteins. Overexpression of these proteins is linked to the abnormal PI3K/AKT/mTOR pathway contributing to increased cancer cell proliferation, a common feature in many cancers including BC [59]. The dysregulation caused by NUDT3-AS4 via DNAm may therefore contribute to increased BC risk.

Most of the identified DNAm sites associated with BC risk independent of genetic effects, and may reflect within-pair differences in exposures to environmental BC risk factors, such as alcohol consumption, exposure to sex hormones [24], and risk factors previously identified in twin studies such as age at first birth, number of children and age at menopause [60] and BMI [61]. Identification of genes related to estrogen signaling (*TDRD1* [62], *SCMH1* [31], *PXDNL* [63], *GNAS* [64] and *RMDN1* [65]) suggests that subtle within-pair differences in hormonal exposures may have resulted in the observed within-pair differences in DNAm patterns associated with future BC diagnosis. In particular, identification of BC–associated DNAm in *SCMH1*, which has been associated with lifetime exposure to sex hormones [31], reinforce this. However, based on the available phenotype data, we could not demonstrate clear differences in hormone exposure (nor other risk factors) between the twin with BC and her healthy sister. However, it is important to recognize that e.g. hormonal exposure due to environmental factors may not be determined by individual factors alone, but rather by the complex combination of multiple factors over time [66], and thereby would be difficult to address using the given phenotype data. There may also be other contributing risk factors, which we have not been able to assess. Investigation of the role of individual risk factors contributing to DNAm changes prior to disease onset requires larger and targeted studies.

All of the observed DNAm differences within the pairs discordant for BC have originated prior to BC diagnosis, and they are mostly driven by environmental effects. We were able to test for direct causation for only 12 of the 212 significant sites as for the rest of the significant CpG sites we could not identify suitable IVs for the MR analysis. For the available IVs only in one CpG site, cg20145695, DNAm could be causally linked to BC. DNAm at cg20145695 increased the risk for ER+ BC and decreased the risk for ER-BC. This CpG site is located within the gene *NXN* coding for nucleoredoxin. Nucleoredoxin interacts with protein flightless-1 homolog [67], which has been demonstrated to enhance genome accessibility at estrogen receptor targets in MCF7 BC cell lines [68]. Changes in *NXN* expression through methylation at cg20145695, could modulate the activity of protein flightless-1 homolog, which impact the accessibility of estrogen targets on the genome. Thereby, the cell’s sensitivity to estrogen signaling could potentially be impacted, distinguishing ER+ from ER-BC, and may explain our finding on *NXN* methylation resulting in opposite risks for ER+ and ER-BC.

Notable strengths of this study were embedded into the study design. The within-pair comparisons account for both known and unknown factors that could confound DNAm analysis, including genotype, age, and familial effects shared by the co-twins that differ for BC diagnosis. The DNAm data used for analysis was collected on average 11 years prior to BC diagnosis, which effectively minimizes the potential confounding of BC as a disease and its treatment on the DNAm increasing the power to detect true associations. However, we saw a limited reproducibility of our findings in the DTS. The DTS is characterized by a considerably smaller sample size, and it thereby likely lacks the statistical power required for robust replication. Previous studies in unrelated cases and controls are prone to genetic confounding, which could explain the lack of replication with our study.

The limited availability of IVs in this study constrained the comprehensive testing for the impact of DNAm on BC risk. However, we did find causal evidence for a single CpG site with its methylation causing the opposite risk for ER+ and ER-BC. This is in line with the notion that the risk for distinct BC subtypes, especially the hormone receptor-positive versus hormone receptor-negative BC are impacted by varying environmental risk factors [69]. In addition to the limited number of suitable IVs for causality analyses, the blood samples for DNAm were collected on average 11 years prior to diagnosis. This extended timespan might be too long to observe strong causal effects of DNAm on BC risk. An alternative explanation could be that DNAm is associated with the risk of the disease through confounding arising from exposures that simultaneously influence both BC risk and DNAm, albeit through different pathways. While understanding the relationship between DNAm and BC risk is crucial for a broader understanding of disease etiology, it is important to note that this limitation does not impact the utility of DNAm as a biomarker for BC risk.

## Conclusion

This study demonstrated the presence of BC–associated DNAm patterns in blood on average 11 years before the actual BC diagnosis, independent of familial factors, likely due to individual environmental exposures. Furthermore, these findings suggest that DNAm could be a promising addition to BC risk assessment toolset for identifying individuals who have a higher likelihood of developing BC. Importantly, our study reveals DNAm at a single CpG site to simultaneously increase the risk for ER+ and decrease the risk for ER-BC. Future studies in larger prospective cohorts are warranted to clarify which environmental factors are most relevant, and increase the risk for BC through DNAm.

## Supporting information

Supplementary_Data_Bode_etal_2024

## List of abbreviations

BC: breast cancer
DTS: Danish Twin Study
DZ: dizygotic
EPIC: Illumina Infinium MethylationEPIC BeadChip
ER+ BC: estrogen receptor positive breast cancer
ER-BC: estrogen receptors negative breast cancer
EWAS: epigenome wide association study
FTC: Finnish Twin Cohort Study
GWAS: genome wide association study
HR: Hazard Ratio
IV: instrumental variable
LSADT: Longitudinal Study of Aging in Danish Twins Study
MADT: Middle Age Danish Twins Study
MR: Mendelian randomization
MZ: monozygotic
OR: Odds Ratio
UKBB: UK Biobank
450K: Infinium HumanMethylation450 BeadChip

## Declarations

### Ethics approval and consent to participate

Informed consent was obtained before the beginning of the FTC studies in 1975, and upon every contact with the study subjects. When clinical investigations were undertaken with sampling of biological material, written informed consent was obtained. Ethics approval of these procedures was provided in multiple studies, the last one on the transfer of biological samples to the THL Biobank by the Hospital District of Helsinki and Uusimaa ethics board in 2018 (#1799/2017). Permission for register linkage to the Finnish Cancer Registry was given by the Finnish Social and Health Data Permit Authority Findata (latest THL/6353/14.06.00/2023).

The two Danish twin studies (LSADT and MADT) were approved by the Regional Committees on Health Research Ethics for Southern Denmark (S-VF-19980072), and written informed consents were obtained from all participants.

### Availability of data and materials

The Finnish Twin Cohort phenotype and methylation data utilized in the current study are deposited in the Biobank of the Finnish Institute for Health and Welfare, Helsinki, Finland. Access to Finnish Cancer Registry data can be applied for as part of the Biobank access procedures. These data are available for use by qualified researchers through a standardized application procedure (https://thl.fi/en/statistics-and-data/data-and-services/research-use-and-data-permits). Survey data and biological samples for the Danish Twin Study are available at the Danish Twin Registry at the Department of Public Health, University of Southern Denmark, Odense, Denmark (https://www.sdu.dk/en/om_sdu/institutter_centre/ist_sundhedstjenesteforsk/centre/dtr/researcher). For more information on the Danish Twin Registry please refer to Pedersen et al. 2019 [36].

### Authors’ contributions

All authors contributed to conceptualization and methodology. HB and LH contributed to the bioinformatics and statistical analyses. HB, MO and JK contributed to the interpretation. HB contributed to writing of the original draft; all authors contributed to reviewing and editing of the manuscript. JH, LH, JK and MO contributed to supervision of the project. All authors read and approved the final manuscript.

## Acknowledgements

The authors thank the participants for their invaluable contribution to the study. The technical staff at the Finnish Twin Cohort study and the Danish Twin Study are acknowledged for their help in collecting the data.

## References

1. Johansson A, Flanagan JM. Epigenome-wide association studies for breast cancer risk and risk factors. Trends Cancer Res. 2017;12:19–28.

2. Massi MC, Dominoni L, Ieva F, Fiorito G. A Deep Survival EWAS approach estimating risk profile based on pre-diagnostic DNA methylation: An application to breast cancer time to diagnosis. PLoS Comput Biol. 2022;18(9):e1009959. Published 2022 Sep 26. doi:10.1371/journal.pcbi.1009959

3. Joo JE, Dowty JG, Milne RL, et al. Heritable DNA methylation marks associated with susceptibility to breast cancer. Nat Commun. 2018;9(1):867. Published 2018 Feb 28. doi:10.1038/s41467-018-03058-6

4. Kresovich JK, Xu Z, O’Brien KM, et al. Blood DNA methylation profiles improve breast cancer prediction. Mol Oncol. 2022;16(1):42–53. doi:10.1002/1878-0261.13087

5. Tuminello S, Zhang Y, Yang L, et al. Global DNA Methylation Profiles in Peripheral Blood of WTC-Exposed Community Members with Breast Cancer. Int J Environ Res Public Health. 2022;19(9):5104. Published 2022 Apr 22. doi:10.3390/ijerph19095104

6. Ennour-Idrissi K, Dragic D, Issa E, et al. DNA Methylation and Breast Cancer Risk: An Epigenome-Wide Study of Normal Breast Tissue and Blood. Cancers (Basel). 2020;12(11):3088. Published 2020 Oct 23. doi:10.3390/cancers12113088

7. Ho PJ, Dorajoo R, Ivanković I, et al. DNA methylation and breast cancer-associated variants. Breast Cancer Res Treat. 2021;188(3):713–727. doi:10.1007/s10549-021-06185-9

8. Shenker NS, Polidoro S, van Veldhoven K, et al. Epigenome-wide association study in the European Prospective Investigation into Cancer and Nutrition (EPIC-Turin) identifies novel genetic loci associated with smoking. Hum Mol Genet. 2013;22(5):843–851. doi:10.1093/hmg/dds488

9. Yao S, Hu Q, Kerns S, et al. Impact of chemotherapy for breast cancer on leukocyte DNA methylation landscape and cognitive function: a prospective study. Clin Epigenetics. 2019;11(1):45. Published 2019 Mar 12. doi:10.1186/s13148-019-0641-1

10. Wong EM, Southey MC, Terry MB. Integrating DNA methylation measures to improve clinical risk assessment: are we there yet? The case of BRCA1 methylation marks to improve clinical risk assessment of breast cancer. Br J Cancer. 2020;122(8):1133–1140. doi:10.1038/s41416-019-0720-2

11. Heyn H, Carmona FJ, Gomez A, et al. DNA methylation profiling in breast cancer discordant identical twins identifies DOK7 as novel epigenetic biomarker. Carcinogenesis. 2013;34(1):102–108. doi:10.1093/carcin/bgs321

12. Scott CM, Wong EM, Joo JE, et al. Genome-wide DNA methylation assessment of ‘BRCA1-like’ early-onset breast cancer: Data from the Australian Breast Cancer Family Registry. Exp Mol Pathol. 2018;105(3):404–410. doi:10.1016/j.yexmp.2018.11.006

13. Anjum S, Fourkala EO, Zikan M, et al. A BRCA1-mutation associated DNA methylation signature in blood cells predicts sporadic breast cancer incidence and survival. Genome Med. 2014;6(6):47. Published 2014 Jun 27. doi:10.1186/gm567

14. Xu Z, Sandler DP, Taylor JA. Blood DNA Methylation and Breast Cancer: A Prospective Case-Cohort Analysis in the Sister Study. J Natl Cancer Inst. 2020;112(1):87–94. doi:10.1093/jnci/djz065

15. Yang Y, Wu L, Shu XO, et al. Genetically Predicted Levels of DNA Methylation Biomarkers and Breast Cancer Risk: Data From 228 951 Women of European Descent. J Natl Cancer Inst. 2020;112(3):295–304. doi:10.1093/jnci/djz109

16. Xiong Z, Yang L, Ao J, et al. A Prognostic Model for Breast Cancer Based on Cancer Incidence-Related DNA Methylation Pattern. Front Genet. 2022;12:814480. Published 2022 Jan 3. doi:10.3389/fgene.2021.814480

17. Xu Z, Bolick SC, DeRoo LA, Weinberg CR, Sandler DP, Taylor JA. Epigenome-wide association study of breast cancer using prospectively collected sister study samples. J Natl Cancer Inst. 2013;105(10):694–700. doi:10.1093/jnci/djt045

18. Parashar S, Cheishvili D, Mahmood N, et al. DNA methylation signatures of breast cancer in peripheral T-cells. BMC Cancer. 2018;18(1):574. Published 2018 May 18. doi:10.1186/s12885-018-4482-7

19. Cappetta M, Fernandez L, Brignoni L, et al. Discovery of novel DNA methylation biomarkers for non-invasive sporadic breast cancer detection in the Latino population. Mol Oncol. 2021;15(2):473–486. doi:10.1002/1878-0261.12842

20. Tang Q, Cheng J, Cao X, Surowy H, Burwinkel B. Blood-based DNA methylation as biomarker for breast cancer: a systematic review. Clin Epigenetics. 2016;8:115. Published 2016 Nov 14. doi:10.1186/s13148-016-0282-6

21. Chung FF, Maldonado SG, Nemc A, et al. Buffy coat signatures of breast cancer risk in a prospective cohort study. Clin Epigenetics. 2023;15(1):102. Published 2023 Jun 12. doi:10.1186/s13148-023-01509-6

22. Pashayan N, Pharoah P. Population-based screening in the era of genomics. Per Med. 2012;9(4):451–455. doi:10.2217/pme.12.40

23. Garcia-Closas M, Gunsoy NB, Chatterjee N. Combined associations of genetic and environmental risk factors: implications for prevention of breast cancer. J Natl Cancer Inst. 2014;106(11):dju305. Published 2014 Nov 12. doi:10.1093/jnci/dju305

24. Obeagu EI, Obeagu GU. Breast cancer: A review of risk factors and diagnosis. Medicine (Baltimore). 2024;103(3):e36905. doi:10.1097/MD.0000000000036905

25. Varela-Rey M, Woodhoo A, Martinez-Chantar ML, Mato JM, Lu SC. Alcohol, DNA methylation, and cancer. Alcohol Res. 2013;35(1):25–35.

26. Mahna D, Puri S, Sharma S. DNA methylation signatures: Biomarkers of drug and alcohol abuse. Mutat Res Rev Mutat Res. 2018;777:19–28. doi:10.1016/j.mrrev.2018.06.002

27. Dragic D, Chang SL, Ennour-Idrissi K, Durocher F, Severi G, Diorio C. Association between alcohol consumption and DNA methylation in blood: a systematic review of observational studies. Epigenomics. 2022;14(12):793–810. doi:10.2217/epi-2022-0055

28. Dragic D, Ennour-Idrissi K, Michaud A, Chang SL, Durocher F, Diorio C. Association Between BMI and DNA Methylation in Blood or Normal Adult Breast Tissue: A Systematic Review. Anticancer Res. 2020;40(4):1797–1808. doi:10.21873/anticanres.14134

29. Chen M, Wong EM, Nguyen TL, et al. DNA methylation-based biological age, genome-wide average DNA methylation, and conventional breast cancer risk factors. Sci Rep. 2019;9(1):15055. Published 2019 Oct 21. doi:10.1038/s41598-019-51475-4

30. Światowy WJ, Drzewiecka H, Kliber M, et al. Physical Activity and DNA Methylation in Humans. Int J Mol Sci. 2021;22(23):12989. Published 2021 Nov 30. doi:10.3390/ijms222312989

31. Johansson A, Palli D, Masala G, et al. Epigenome-wide association study for lifetime estrogen exposure identifies an epigenetic signature associated with breast cancer risk. Clin Epigenetics. 2019;11(1):66. Published 2019 Apr 30. doi:10.1186/s13148-019-0664-7

32. Levine ME, Lu AT, Chen BH, et al. Menopause accelerates biological aging. Proc Natl Acad Sci U S A. 2016;113(33):9327–9332. doi:10.1073/pnas.1604558113

33. Kresovich JK, Xu Z, O’Brien KM, Weinberg CR, Sandler DP, Taylor JA. Methylation-Based Biological Age and Breast Cancer Risk. J Natl Cancer Inst. 2019;111(10):1051–1058. doi:10.1093/jnci/djz020

34. Rose RJ, Latvala A, Silventoinen K, Kaprio J. Alcohol consumption at age 18-25 and number of children at a 33-year follow-up: Individual and within-pair analyses of Finnish twins. Alcohol Clin Exp Res. 2022;46(8):1552–1564. doi:10.1111/acer.14886

35. Jetté M, Sidney K, Blümchen G. Metabolic equivalents (METS) in exercise testing, exercise prescription, and evaluation of functional capacity. Clin Cardiol. 1990;13(8):555–565. doi:10.1002/clc.4960130809

36. Pedersen DA, Larsen LA, Nygaard M, et al. The Danish Twin Registry: An Updated Overview. Twin Res Hum Genet. 2019;22(6):499–507. doi:10.1017/thg.2019.72

37. Skytthe A, Harris JR, Czene K, et al. Cancer Incidence and Mortality in 260,000 Nordic Twins With 30,000 Prospective Cancers. Twin Res Hum Genet. 2019;22(2):99–107. doi:10.1017/thg.2019.10

38. Harris JR, Hjelmborg J, Adami HO, et al. The Nordic Twin Study on Cancer - NorTwinCan. Twin Res Hum Genet. 2019;22(6):817–823. doi:10.1017/thg.2019.71

39. Min JL, Hemani G, Davey Smith G, Relton C, Suderman M. Meffil: efficient normalization and analysis of very large DNA methylation datasets. Bioinformatics. 2018;34(23):3983–3989. doi:10.1093/bioinformatics/bty476

40. Zhou W, Laird PW, Shen H. Comprehensive characterization, annotation and innovative use of Infinium DNA methylation BeadChip probes. Nucleic Acids Res. 2017;45(4):e22. doi:10.1093/nar/gkw967

41. Inkster AM, Wong MT, Matthews AM, Brown CJ, Robinson WP. Who’s afraid of the X? Incorporating the X and Y chromosomes into the analysis of DNA methylation array data. Epigenetics Chromatin. 2023;16(1):1. Published 2023 Jan 7. doi:10.1186/s13072-022-00477-0

42. Feil R, Khosla S. Genomic imprinting in mammals: an interplay between chromatin and DNA methylation?. Trends Genet. 1999;15(11):431–435. doi:10.1016/s0168-9525(99)01822-3

43. Teschendorff AE, Marabita F, Lechner M, et al. A beta-mixture quantile normalization method for correcting probe design bias in Illumina Infinium 450 k DNA methylation data. Bioinformatics. 2013;29(2):189–196. doi:10.1093/bioinformatics/bts680

44. Pidsley R, Y Wong CC, Volta M, Lunnon K, Mill J, Schalkwyk LC. A data-driven approach to preprocessing Illumina 450K methylation array data. BMC Genomics. 2013;14:293. Published 2013 May 1. doi:10.1186/1471-2164-14-293

45. van Iterson M, Tobi EW, Slieker RC, et al. MethylAid: visual and interactive quality control of large Illumina 450k datasets. Bioinformatics. 2014;30(23):3435–3437. doi:10.1093/bioinformatics/btu566

46. Aryee MJ, Jaffe AE, Corrada-Bravo H, et al. Minfi: a flexible and comprehensive Bioconductor package for the analysis of Infinium DNA methylation microarrays. Bioinformatics. 2014;30(10):1363–1369. doi:10.1093/bioinformatics/btu049

47. Soerensen M, Hozakowska-Roszkowska DM, Nygaard M, et al. A Genome-Wide Integrative Association Study of DNA Methylation and Gene Expression Data and Later Life Cognitive Functioning in Monozygotic Twins. Front Neurosci. 2020;14:233. Published 2020 Apr 9. doi:10.3389/fnins.2020.00233

48. Therneau T (2024). A Package for Survival Analysis in R. R package version 3.5-8, https://CRAN.R-project.org/package=survival.

49. VanderWeele TJ, Ding P. Sensitivity Analysis in Observational Research: Introducing the E-Value. Ann Intern Med. 2017;167(4):268–274. doi:10.7326/M16-2607

50. Korhonen T, Hjelmborg J, Harris JR, et al. Cancer in twin pairs discordant for smoking: The Nordic Twin Study of Cancer. Int J Cancer. 2022;151(1):33–43. doi:10.1002/ijc.33963

51. Xu Z, Xie C, Taylor JA, Niu L. ipDMR: identification of differentially methylated regions with interval P-values. Bioinformatics. 2021;37(5):711–713. doi:10.1093/bioinformatics/btaa732

52. Xu Z, Niu L, Taylor JA. The ENmix DNA methylation analysis pipeline for Illumina BeadChip and comparisons with seven other preprocessing pipelines. Clin Epigenetics. 2021;13(1):216. Published 2021 Dec 9. doi:10.1186/s13148-021-01207-1

53. Villicaña S, Castillo-Fernandez J, Hannon E, et al. Genetic impacts on DNA methylation help elucidate regulatory genomic processes. Genome Biol. 2023;24(1):176. Published 2023 Jul 31. doi:10.1186/s13059-023-03011-x

54. Elsworth B, Lyon M, Alexander T, Liu Y, Matthews P, Hallett J, Bates P, Palmer T, Haberland V, Smith GD, Zheng J, Haycock P, Gaunt TR, & Hemani G. The MRC IEU OpenGWAS data infrastructure. bioRxiv. 2020. 10.1101/2020.08.10.244293

55. Burrows K, Bull CJ, Dudding T, Gormley M, Robinson T, Tan V, et al. Genome-wide Association Study of Cancer Risk in UK Biobank. University of Bristol, UK; 2021. doi:10.5523/bris.aed0u12w0ede20olb0m77p4b9

56. Kamat MA, Blackshaw JA, Young R, et al. PhenoScanner V2: an expanded tool for searching human genotype-phenotype associations. Bioinformatics. 2019;35(22):4851–4853. doi:10.1093/bioinformatics/btz469

57. Hemani G, Tilling K, Davey Smith G. Orienting the causal relationship between imprecisely measured traits using GWAS summary data [published correction appears in PLoS Genet. 2017 Dec 29;13(12):e1007149]. PLoS Genet. 2017;13(11):e1007081. Published 2017 Nov 17. doi:10.1371/journal.pgen.1007081

58. Hemani G, Zheng J, Elsworth B, et al. The MR-Base platform supports systematic causal inference across the human phenome. Elife. 2018;7:e34408. Published 2018 May 30. doi:10.7554/eLife.34408

59. Hao Q, Wang P, Dutta P, et al. Comp34 displays potent preclinical antitumor efficacy in triple-negative breast cancer via inhibition of NUDT3-AS4, a novel oncogenic long noncoding RNA. Cell Death Dis. 2020;11(12):1052. Published 2020 Dec 11. doi:10.1038/s41419-020-03235-w

60. Hamilton AS, Mack TM. Puberty and genetic susceptibility to breast cancer in a case-control study in twins. N Engl J Med. 2003;348(23):2313–2322. doi:10.1056/NEJMoa021293

61. Lundqvist E, Kaprio J, Verkasalo PK, et al. Co-twin control and cohort analyses of body mass index and height in relation to breast, prostate, ovarian, corpus uteri, colon and rectal cancer among Swedish and Finnish twins. Int J Cancer. 2007;121(4):810–818. doi:10.1002/ijc.22746

62. Kacprzyk LA, Laible M, Andrasiuk T, et al. ERG induces epigenetic activation of Tudor domain-containing protein 1 (TDRD1) in ERG rearrangement-positive prostate cancer. PLoS One. 2013;8(3):e59976. doi:10.1371/journal.pone.0059976

63. Lu Y, Li J, Cheng J, Lubahn DB. Messenger RNA profile analysis deciphers new Esrrb responsive genes in prostate cancer cells. BMC Mol Biol. 2015;16:21. Published 2015 Dec 1. doi:10.1186/s12867-015-0049-1

64. Pham LT, Yamanaka K, Miyamoto Y, Waki H, Gouraud SSS. Estradiol-dependent gene expression profile in the amygdala of young ovariectomized spontaneously hypertensive rats. Physiol Genomics. 2022;54(3):99–114. doi:10.1152/physiolgenomics.00082.2021

65. Yazdanpanah N, Jumentier B, Yazdanpanah M, Ong KK, Perry JRB, Manousaki D. Mendelian randomization identifies circulating proteins as biomarkers for age at menarche and age at natural menopause. Commun Biol. 2024;7(1):47. Published 2024 Jan 6. doi:10.1038/s42003-023-05737-7

66. Clusan L, Ferrière F, Flouriot G, Pakdel F. A Basic Review on Estrogen Receptor Signaling Pathways in Breast Cancer. Int J Mol Sci. 2023;24(7):6834. Published 2023 Apr 6. doi:10.3390/ijms24076834

67. Idelfonso-García OG, Alarcón-Sánchez BR, Vásquez-Garzón VR, et al. Is Nucleoredoxin a Master Regulator of Cellular Redox Homeostasis? Its Implication in Different Pathologies. Antioxidants (Basel). 2022;11(4):670. Published 2022 Mar 30. doi:10.3390/antiox11040670

68. Jeong KW. Flightless I (Drosophila) homolog facilitates chromatin accessibility of the estrogen receptor α target genes in MCF-7 breast cancer cells. Biochem Biophys Res Commun. 2014;446(2):608–613. doi:10.1016/j.bbrc.2014.03.011

69. Waks AG, Winer EP. Breast Cancer Treatment: A Review. JAMA. 2019;321(3):288–300. doi:10.1001/jama.2018.19323

