## Supplementary_Figure_1 for "Pre-diagnosis blood DNA methylation profiling of twin pairs discordant for breast cancer points to the importance of environmental risk"

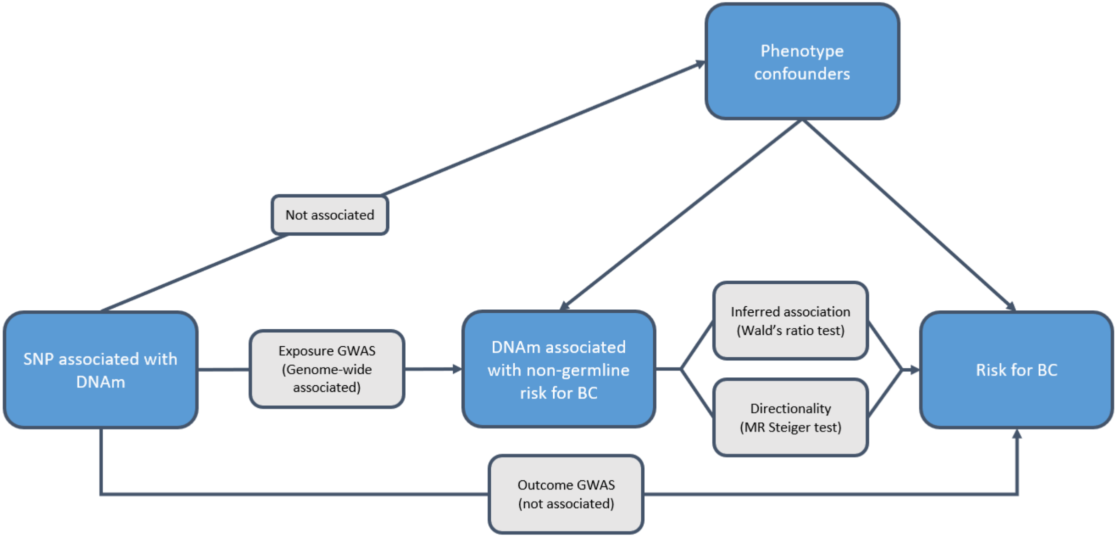


Supplementary Figure 1: Schematic representation of the MR analysis. SNPs are tested for association with phenotype confounders, BC and DNAm at given meQTLs (identified CpG sites in Model 1) to only select appropriate instrumental variables. The causal inference of DNAm on risk for BC (overall, ER+ and ER- BC) is tested by Wald's Ratio using genetic associations of the instrumental variables with the exposure and the outcome. Directionality from DNAm on risk for BC was assessed using the MR Steiger test.
